# Does place connectivity moderate the association between concentrated disadvantage and COVID-19 fatality in the United States?

**DOI:** 10.1101/2022.06.06.22276053

**Authors:** Fengrui Jing, Zhenlong Li, Shan Qiao, Jiajia Zhang, Bankole Olatosi, Xiaoming Li

## Abstract

Concentrated disadvantaged areas have been disproportionately affected by COVID-19 outbreak in the United States (US). Meanwhile, highly connected areas may contribute to higher human movement, leading to higher COVID-19 cases and deaths. This study examined whether place connectivity moderated the association between concentrated disadvantage and COVID-19 fatality. Using COVID-19 fatality over four time periods, we performed mixed-effect negative binomial regressions to examine the association between concentrated disadvantage, Twitter-based place connectivity, and county-level COVID-19 fatality, considering potential state-level variations. Results revealed that concentrated disadvantage was significantly associated with an increased COVID-19 fatality. More importantly, moderation analysis suggested that place connectivity significantly exacerbated the harmful effect of concentrated disadvantage on COVID-19 fatality, and this significant moderation effect increased over time. In response to COVID-19 and other future infectious disease outbreaks, policymakers are encouraged to focus on the disadvantaged areas that are highly connected to provide additional pharmacological and non-pharmacological intervention policies.

## Introduction

By 2022, the Coronavirus Disease 2019 (COVID-19) pandemic has entered its third year. To curb the transmission and deaths from COVID-19, besides pharmacological interventions, the government has implemented a range of non-pharmacological interventions (NPIs) such as social distancing, face mask orders, remote work, travel restrictions, and lockdowns. The effects of non-pharmaceutical policies vary by group and have led to a range of health disparities. Further research on health disparities in COVID-19 outbreaks is necessary to guide the prevention and control of COVID-19 and related infectious diseases in the future.

Several studies have shown that areas with concentrations of socioeconomically disadvantaged people are strongly associated with more health problems, including disproportionately high transmission and deaths of COVID-19 (Levy et al., 2022; Pierce, & Jacob, 2021; Samuels-Kalow et al., 2021). Disadvantaged populations live in house conditions that do not allow for effective voluntary isolation and quarantine (Duque, 2020; Khanijahani, & Tomassoni, 2022), and suffer from poorer health status (Barber et al., 2021). Another significant factor could be the higher population mobility of disadvantaged populations. During the pandemic, especially under strict policies such as social distancing and mobility restrictions, disadvantaged populations have higher mobility since most of them are engaged in essential work (Bonaccorsi, 2020; Sy et al., 2021). Thus, the high mobility in disadvantaged areas may contribute to COVID-19 outbreaks in these areas. However, few studies have considered the role of connectivity in disadvantaged areas, that is, the interaction effect of concentrated disadvantage and connectivity on COVID-19. As a relatively stable characteristic of an area/neighborhood, high connectivity can foster population mobility within disadvantaged areas and thus may further exacerbate the negative impacts of concentrated disadvantages on COVID fatality.

In this paper, we use Twitter-based place connectivity to test whether place connectivity contributes to the adverse consequences of living in concentrated disadvantaged areas on COVID-19 fatality. The findings may provide insight into the relationship between concentrated disadvantage and COVID-19 fatality.

### Concentrated disadvantage and COVID-19 fatality

Concentrated disadvantage, also known as neighborhood disadvantage, or deprivation index in some cases, refers to areas with a high proportion of people with low socioeconomic status. Concentrated disadvantaged areas aggregate groups such as low-income earners, welfare recipients, and single households (Sampson, Raudenbush, & Earls, 1997; Sampson, Sharkey, & Raudenbush, 2008), and some may also include minority groups (Lee, Maume, & Ousey, 2003). These groups face higher levels of health risks (Barber et al., 2021) and crime risks (Lee, Maume, & Ousey, 2003; Wang, & Arnold, 2008). Many studies have revealed the significant relationships between concentrated disadvantages and differential forms of health inequalities, such as the increased risk of breast cancer (Barber et al., 2021; DeGuzman, 2017), increased incidence of lung cancer (Adie et al., 2020), diabetes and cholesterol control (Durfey et al., 2019), mental health (Kim, 2010).

The distribution of COVID-19 transmission and fatality shows spatial disparities. Concentrated disadvantaged areas are more likely to suffer disproportionate COVID-19 infection (Levy et al., 2022; Yellow Horse, Yang, & Huyser, 2022), deaths (Pierce, & Jacob, 2021), and fatality (Holmes et al., 2020; Sen-Crowe et al., 2021a). At the individual level, these areas are concentrated with residents of poorer socioeconomic status, who are in poorer health and are more likely to be essential workers in professions such as grocery delivery, truck drivers, and cleaners (Ghilarducci, & Farmand, 2020; Huyser, Yang, & Horse, 2021; Khanijahani, & Tomassoni, 2022). Most of these jobs are difficult to perform remotely and lack the conditions to maintain social distancing. Meanwhile, these groups may use public transportation more frequently, such as a study in New York City found that areas with low-income people, essential workers, and non-white populations had more mobility extracted from subway data during the pandemic (Sy et al., 2021). Disadvantaged populations also live in mostly poor house conditions, with many living together and without good post-infection isolation (Boateng et al., 2021; Truong & Asare, 2021). These factors of physical status, work environment, commuting patterns, and house conditions contribute to a higher risk of exposure to COVID-19 and the increased likelihood of COVID-19 infection and fatality in socioeconomically disadvantaged populations. Conversely, areas with high socioeconomic status may concentrate residents who are in good health status, can work remotely, and live in house conditions that allow isolation from infected family members, which may reduce the risk of infection and fatality.

Exploration of the association between concentrated disadvantage and COVID-19 is generally at the neighborhood (Jr, 2021), zip code (Yellow Horse, Yang, & Huyser, 2022), or county scale (Kranjac, & Kranjac, 2021), as well as community-based county-scale indicators (Khanijahani, & Tomassoni, 2022). However, to date, most studies have focused on the incidence and mortality in the early stages of COVID-19 outbreaks, and fewer studies consider different periods (Tokey, 2021; Lee, 2021). The fact is that COVID-19 is constantly mutating and spreading, and the impact of concentrated disadvantage on virus spread and fatality may vary during different periods. Just like some studies have demonstrated that human mobility and education level affect COVID-19 infection rate at different stages (Tokey, 2021).

### Moderation effects of connectivity

The connectivity of a place can be described as the strength of a connection between a place and one or more places, and this connection is generally manifested in terms of the road, train, air, and social media, among others. Unlike direct population movements, connectivity is more stable, as it is closely related to geographical location, transportation facilities, and other related static factors. The greater the connectivity between areas, the higher level of population mobility between these areas. Given human mobility is a significant predictor of COVID-19 incidence (Zeng et al., 2021), higher connectivity could be associated with a higher risk of exposure, and greater risk of COVID-19 infection. Several studies have found that air connectivity (Sun, Wandelt, & Zhang, 2021), high-speed train connectivity (Zhang, Zhang, & Wang, 2020; Zhu, & Guo, 2021), road connectivity (Cuadros et al., 2020), and Twitter-based place connectivity (Li et al., 2021) are associated with the initial outbreak of COVID-19. Particularly, Twitter-based connectivity, representing the extent to which a place shares the same users with other places, gives a comprehensive measure of the degree of connectivity in all aspects of transportation in that place, which can be a more direct proxy for population mobility and exposure risk (Li et al., 2021).

There are few studies with mixed results regarding the association between connectivity and COVID-19 clinical consequences (including fatality). Some studies have shown a significant association between air connectivity index and increased death (Fountoulakis et al., 2020) and death risk (Correa-Agudelo et al., 2020) in early-stage, while another suggested that pedestrian-oriented street connectivity is associated with lower COVID-19 death rates, because residents in this built environment may have higher physical activity and lower levels of obesity and chronic disease (Wali, & Frank, 2021). To date, no studies have directly analyzed the relationship between place connectivity and COVID-19 fatality. Given fatality is more influenced by age structure and the quality of the healthcare system (Moosa, & Khatatbeh, 2021), the association between connectivity and COVID-19 fatality may be indirect or connectivity may moderate the impact of various factors such as health infrastructure in the area, access to health services, and pre-existing health conditions of a population on COVID-19 clinical outcomes.

As many studies have confirmed (Ghilarducci, & Farmand, 2020; Khanijahani, & Tomassoni, 2022), people living in high concentrated disadvantaged areas may have higher needs to travel because most of them are essential workers and have limited resources to support remote working. In this case, if the area is also highly connected, these people may be more likely to take advantage of the convenient connectivity conditions (e.g., transportation) to go to work. Under the implementation of NPIs like lockdown during the pandemic, a high connectivity place with a concentration of disadvantaged groups may have higher mobility compared to other high connectivity places without a concentration of disadvantaged groups. Higher mobility is associated with higher rates of infection (Sun, Wandelt, & Zhang, 2021; Zhang, Zhang, & Wang, 2020). For people living in disadvantaged areas, a higher infection rate is usually linked with higher fatality given their poor pre-existing health conditions (Ross, & Mirowsky, 2001) and barriers to access to healthcare services (Kirby, & Kaneda, 2005; Peters et al., 2008). Therefore, a hypothesis is that connectivity may amplify the negative impacts of concentrated disadvantage on COVID-19 fatality.

Although several studies have demonstrated the association between mobility and concentrated disadvantage (Benitez, J., Courtemanche, & Yelowitz, 2020; Sy et al., 2021), and the effects of human mobility and concentrated disadvantage on COVID-19, respectively (Samuels-Kalow et al., 2021; Zhang, Zhang, & Wang, 2020), to the best of our knowledge, no study has yet tested the moderation effect of connectivity on the association between concentrated disadvantage and COVID-19 fatality. If this moderation is confirmed, it is meaningful for the prevention and control of COVID-19 and similar infectious diseases and would further support the significance of the two factors (i.e, concentrated disadvantage and connectivity) in the NPIs during the pandemic.

### The present study

This paper proposes that place connectivity can intensify the harmful effects of county-level concentrated disadvantage on county-level COVID-19 fatality. If a county with a concentration of disadvantaged populations is also a highly connected county, the disadvantaged group will have higher mobility through place connectivity and a greater probability of exposure to the virus, which may contribute to the deleterious effect of concentrated disadvantage on COVID-19 fatality. If so, this may help further explain why concentrated disadvantage is associated with a high COVID-19 fatality. Furthermore, since the pandemic has been prevalent for a long time, and travel restriction policies have changed across time, place connectivity may not contribute to population movement in the same way, such that the effect of place connectivity on concentrated disadvantage and COVID-19 fatality may be varied.

In this paper, we use Twitter data to measure place connectivity. Twitter-based place connectivity is based on real Twitter users and is comprehensive connectivity, as previous studies have noted that it reflects connectivity not only in terms of transportation, but also in terms of social networks, geography, and socioeconomics (Li et al., 2021). Meanwhile, given the close association with these relatively static factors, place connectivity is a stable factor across years before the pandemic (Li et al., 2021). This study uses historical place connectivity to analyze its relationship with current COVID-19 fatality, which will be useful in guiding the role place connectivity may play in future infectious disease prevention and control.

Specifically, we present the following hypotheses:

H1: Concentrated disadvantage is associated with higher COVID-19 fatality.

H2: The association between concentrated disadvantage and COVID-19 fatality is stronger in counties of high place connectivity compared to counties of low place connectivity.

H3: The moderation effect of place connectivity may vary along with the period of the pandemic.

## Methods

### Data sources and study area

We obtained the county-level data for confirmed COVID-19 cases and deaths from the start of the outbreak on January 21, 2020, to December 1, 2022, in the contiguous US from the New York Times (https://github.com/nytimes/covid-19-data). This data was initially collected from the Center for Disease Control and Prevention (CDC), multilevel health departments, and other related sources. It has been used in numerous studies (Janke et al., 2021; Khazanchi et al., 2020). For data on county-level socioeconomic variables, we used the American Community Survey (ACS) 5-year estimates (2015-2019). Data from ACS 1-year or 3-year were not used because these data are limited to areas with populations over 20,000, and the current study intended to ensure data availability for smaller counties with populations less than 20,000 (Khanijahani & Tomassoni, 2022). Twitter is one of the most popular social media platforms in the US and a very prevalent source of geospatial social media data in academia. We used place connectivity extracted from Twitter in 2018 and 2019 (Li et al., 2021).

The study area is the contiguous US, with 3,091 counties. Omitted counties are due for two reasons. First, Twitter-based connectivity data covered 3,113 counties in the contiguous US. Second, since the subsequent empirical analysis used fatality for four time periods, we removed those counties with 0 cumulative COVID-19 cases in each time period, respectively. The spatial unit of this study is the county level.

### Measures

#### COVID-19 fatality

COVID-19 fatality is the outcome variable, which is the cumulative COVID-19 deaths divided by the cumulative COVID-19 cases up to a time period. According to a report from CDC (https://www.cdc.gov/mmwr/volumes/71/wr/mm7104e4.htm#:∼:text=The%20SARS%2DCoV%2D2%20B,associated%20ED%20visits%20and%20hospitalizations), there are three periods of high-COVID-19 transmission: December 1, 2020–February 28, 2021 (winter period); July 15, 2021–October 31, 2021 (Delta predominance); and December 19, 2021–January 15, 2022 (Omicron predominance). Correspondingly, the remaining were three normal-COVID-19 transmission periods. Because Omicron is less lethal and not quite the same as previous virus variants, we then selected data up to October 31, 2021, to test our hypotheses.

More specifically, this study includes models for four time periods, based on fatality data up to December 1, 2020 (period 1), up to February 28, 2021 (period 2), up to July 15, 2021 (period 3), and up to October 31, 2021 (period 4), respectively. We did not use single-period data (e.g. 12/1/2020-2/28/2021) to calculate fatality because the death population for a single period did not always belong to the cases in that single period.

#### Concentrated disadvantage

Data on concentrated disadvantage in each county were retrieved from the 5-year estimate American Community Survey (2015-2019). We first defined the concentrated disadvantage variable following previous studies (Sampson, Raudenbush, & Earls, 1997; Lee et al., 2003; Khanijahani & Tomassoni, 2022). We then performed the principal component analysis of five variables and identified these variables loading onto a single factor that accounted for 58.24 % of the observed variation with high reliability (Cronbach’s Alpha α = 0.762). Concentrated disadvantages include five items, the civilian unemployment rate; the percentage of female-headed families; the percentage of the population over the age of 25 that are high school dropouts; the percentage of households with an annual income < $15,000; the percentage of households receiving public assistance. These items are combined into an index by taking the average of their z-scores. Higher values refer to a more concentrated disadvantage index.

#### Place connectivity

Connectivity in this study is assessed by place connectivity index (PCI) extracted through geotagged Tweets (Li et al., 2021). PCI refers to the normalized number of shared Twitter users between the two places in a year (Equation 1). Unlike real-time population movement between places, PCI provides a relatively stable measure of the strength of connectivity between two places through spatial interaction. In this study, we aggregated the PCI values of a county with all other connected counties as the place connectivity of the county (Equation 2), which is the moderator variable of this study. Place connectivity was calculated for 2019 and 2018, separately.

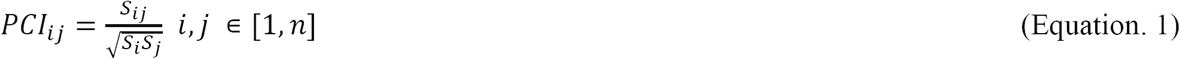

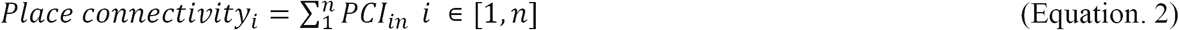

In the equations, *S*_*i*_ is the number of unique Twitter users in county *i* within time T; *S*_*j*_ is the number of unique Twitter users in county *j* within time T; *S*_*i j*_ is the number of shared users between county *i* and *j* within time T; and *n* is the number of counties in the study area.

#### Covariates

We also controlled for other main variables that may affect COVID-19 fatality. For socioeconomic aspects, population density, and uninsured population were included to account for their potential impacts (Gupta et al., 2020; Yellow Horse, Yang, & Huyser, 2022). For demographics, the percentage of the population aged 65 and older was included to control for high-risk groups with high COVID-19 fatality (Yellow Horse, Yang, & Huyser, 2022). The percentage of black or African Americans was included to control for the impact of racial factors on COVID-19 (Khanijahani & Tomassoni, 2022). In addition, the percentage of public transportation commuting was included to account for the impact of public transportation on COVID-19 transmission (Zeng et al., 2021); the percentage of ICU beds was also included to adjust for possible variation in COVID-19 deaths due to differences in availability of healthcare services (Sen-Crowe et al., 2021b). For geographic factors, the geographical census region (Northeast, Midwest, South, and West) was to adjust for the potential impacts of environment-related factors (e.g., temperature and humidity) on the spread and severity of COVID-19 (Khanijahani & Tomassoni, 2022). Core-based statistical area (CBSA) regions were included to adjust for the potential impact of urban and rural factors on COVID-19 fatality (Ahmed et al., 2020; Iyanda, Boakye, & Oppong, 2020). Just as some studies have used geographically weighted models to examine COVID-19 transmission and mortality to control for the role of spatially autocorrelated factors (Yellow Horse, Yang, & Huyser, 2022), this study incorporates spatially lagged fatality to ensure that the model can reduce the effect of spatially autocorrelated factors. Detailly, Moran’ *I* values of county-level COVID-19 fatality in the US were statistically significantly greater than 0 for different time periods, indicating that COVID-19 fatality was spatially correlated, we then included spatially lagged fatality (i.e., COVID-19 fatality in surrounding adjacent counties) to account for spatial autocorrelation of fatality. Table 1 summarized all the key variables in this study.

**Table 1.** Definitions of key variables

| Variables | Definitions |
| --- | --- |
| ***Outcome*** |  |
| COVID-19 fatality | The cumulative COVID-19 deaths divided by the cumulative COVID-19 cases up to a time period |
| ***County-level predictors*** |  |
| ***Demographic characteristics*** |  |
| Concentrated disadvantage | For counties with high percentages of residents of low socioeconomic status (welfare receipt, poverty, unemployment, uneducated, female-headed households) |
| Place connectivity | The total PCI value of the county |
| Spatially lagged fatality | The average fatality in the surrounding counties |
| Population density (per square miles) | The rate of total population to land area in the county |
| % of population aged 65 + | Proportion of population aged 65 + to total population |
| % of no health insurance coverage | Proportion of population with no health insurance coverage to total population |
| % of black or African Americans | Proportion of population of black or African descent to total population |
| % of workers 16 years and over who commute by public transportation | Proportion of population of workers 16 years and over who commute by public transportation |
| ICU bed per 100,000 people | The rate of ICU beds to total population in the county multiplied by 100,000 |
| Core-based statistical area (CBSA) |  |
| No-CBSA | 0 |
| Micropolitan statistical area | 1 |
| Metropolitan statistical area | 2 |
| Region |  |
| Northeast | 0 |
| Midwest | 1 |
| South | 2 |
| West | 3 |

### Statistical Analysis

The count data for COVID-19 deaths were highly right-skewed and overdispersed. As Poisson regression could not capture overdispersion, the negative binomial model is more appropriate. Considering the differential impact of COVID-19 fatality rates by state-level policies such as social distance, face masks, and home orders, we further selected a mixed-effects negative binomial regression model to account for state-level random effects on COVID-19 fatality at the county level (Khanijahani, & Tomassoni, 2022). In each model, to calculate the fatality, the number of COVID-19 deaths was the dependent variable, and the number of COVID-19 cases was the offset term. To avoid numerical singularities in estimating the models, we log-transformed certain variables (population density, place connectivity, and spatially lagged fatality) to ensure accurate analytical results. Before the regression analysis, we used Pearson correlation and VIF analysis to examine possible co-collinearity. The results show that no significant co-collinearity exists between variables (VIF values less than 4). We performed statistical analyses in Stata SE version 15.

## Results

### Descriptive statistics and spatial characteristics

Table 2 presents the descriptive statistics of the variables. By October 31, 2021 (period 4), the county-level average fatality is 0.018. The score range for concentrated disadvantage was from -1.442 to 4.916, with a mean value of -0.073. The average place connectivity in 2019 was 5,634.516, varying from 1,167.885 to 24,065.32. The statistics of other variables were shown in Table 2.

**Table 2.**
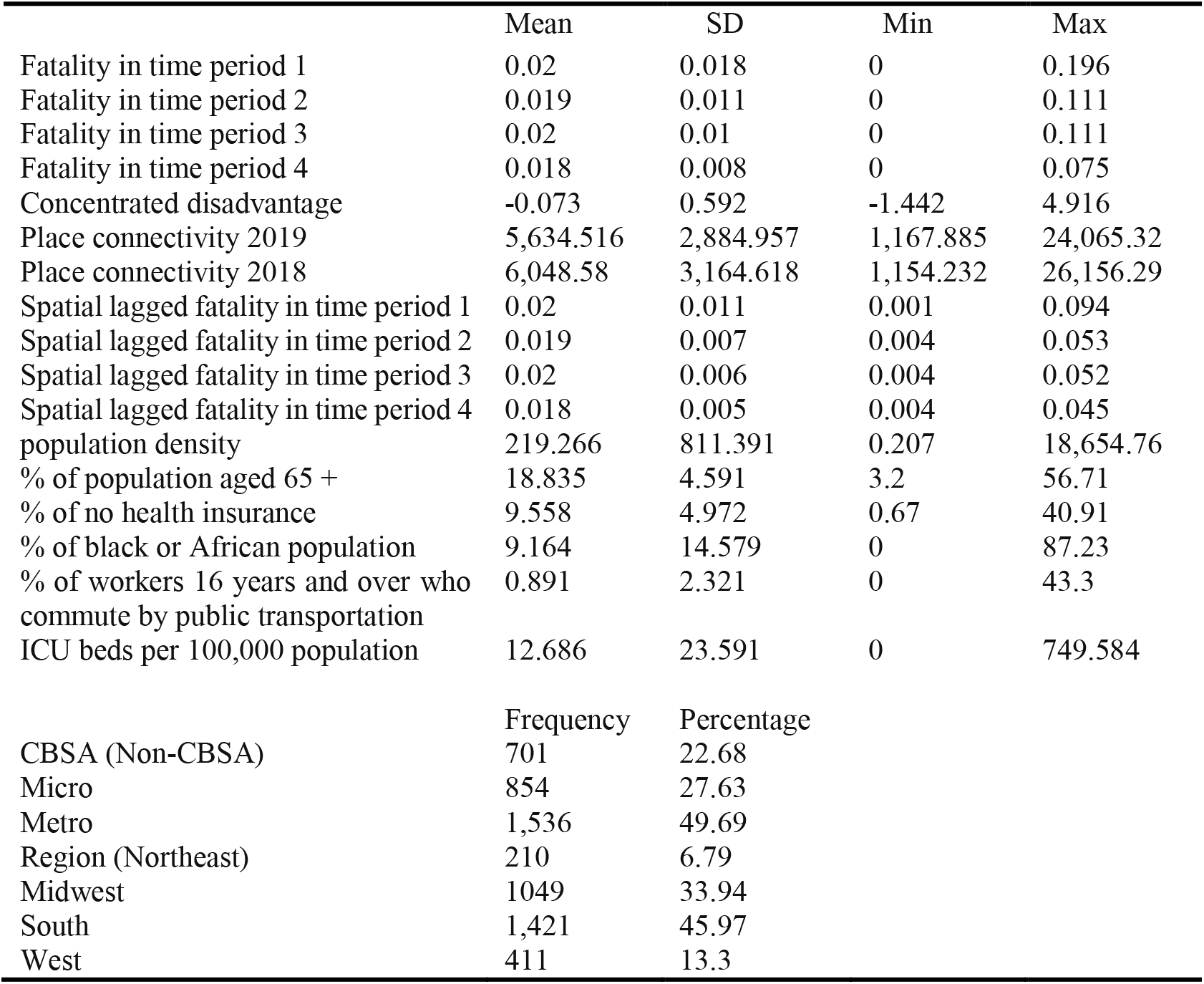
The descriptive statistics of the variables

|  | Mean | SD | Min | Max |
| --- | --- | --- | --- | --- |
| Fatality in time period 1 | 0.02 | 0.018 | 0 | 0.196 |
| Fatality in time period 2 | 0.019 | 0.011 | 0 | 0.111 |
| Fatality in time period 3 | 0.02 | 0.01 | 0 | 0.111 |
| Fatality in time period 4 | 0.018 | 0.008 | 0 | 0.075 |
| Concentrated disadvantage | -0.073 | 0.592 | -1.442 | 4.916 |
| Place connectivity 2019 | 5,634.516 | 2,884.957 | 1,167.885 | 24,065.32 |
| Place connectivity 2018 | 6,048.58 | 3,164.618 | 1,154.232 | 26,156.29 |
| Spatial lagged fatality in time period 1 | 0.02 | 0.011 | 0.001 | 0.094 |
| Spatial lagged fatality in time period 2 | 0.019 | 0.007 | 0.004 | 0.053 |
| Spatial lagged fatality in time period 3 | 0.02 | 0.006 | 0.004 | 0.052 |
| Spatial lagged fatality in time period 4 | 0.018 | 0.005 | 0.004 | 0.045 |
| population density | 219.266 | 811.391 | 0.207 | 18,654.76 |
| % of population aged 65 + | 18.835 | 4.591 | 3.2 | 56.71 |
| % of no health insurance | 9.558 | 4.972 | 0.67 | 40.91 |
| % of black or African population | 9.164 | 14.579 | 0 | 87.23 |
| % of workers 16 years and over who commute by public transportation | 0.891 | 2.321 | 0 | 43.3 |
| ICU beds per 100,000 population | 12.686 | 23.591 | 0 | 749.584 |
|  | Frequency | Percentage |  |  |
| CBSA (Non-CBSA) | 701 | 22.68 |  |  |
| Micro | 854 | 27.63 |  |  |
| Metro | 1,536 | 49.69 |  |  |
| Region (Northeast) | 210 | 6.79 |  |  |
| Midwest | 1049 | 33.94 |  |  |
| South | 1,421 | 45.97 |  |  |
| West | 411 | 13.3 |  |  |

The geospatial distribution of fatality in the contiguous US shows that the high fatality area was widely distributed and tends to be concentrated in the South (Figure 1). Economically developed regions like California did not show an excessive fatality. From the four time periods, the areas of high mortality changed over time. Initially, hotspots were in the Northeast and Southwest, and then gradually spread to the interior and surrounding regions. This may be related to coronavirus transmission, as the outbreak first occurred in the metropolitan areas of the east and west coasts. There was a trend of high correlation between fatality and spatially lagged fatality in each county, which corroborates the infectious character of the virus.

**Figure 1.**
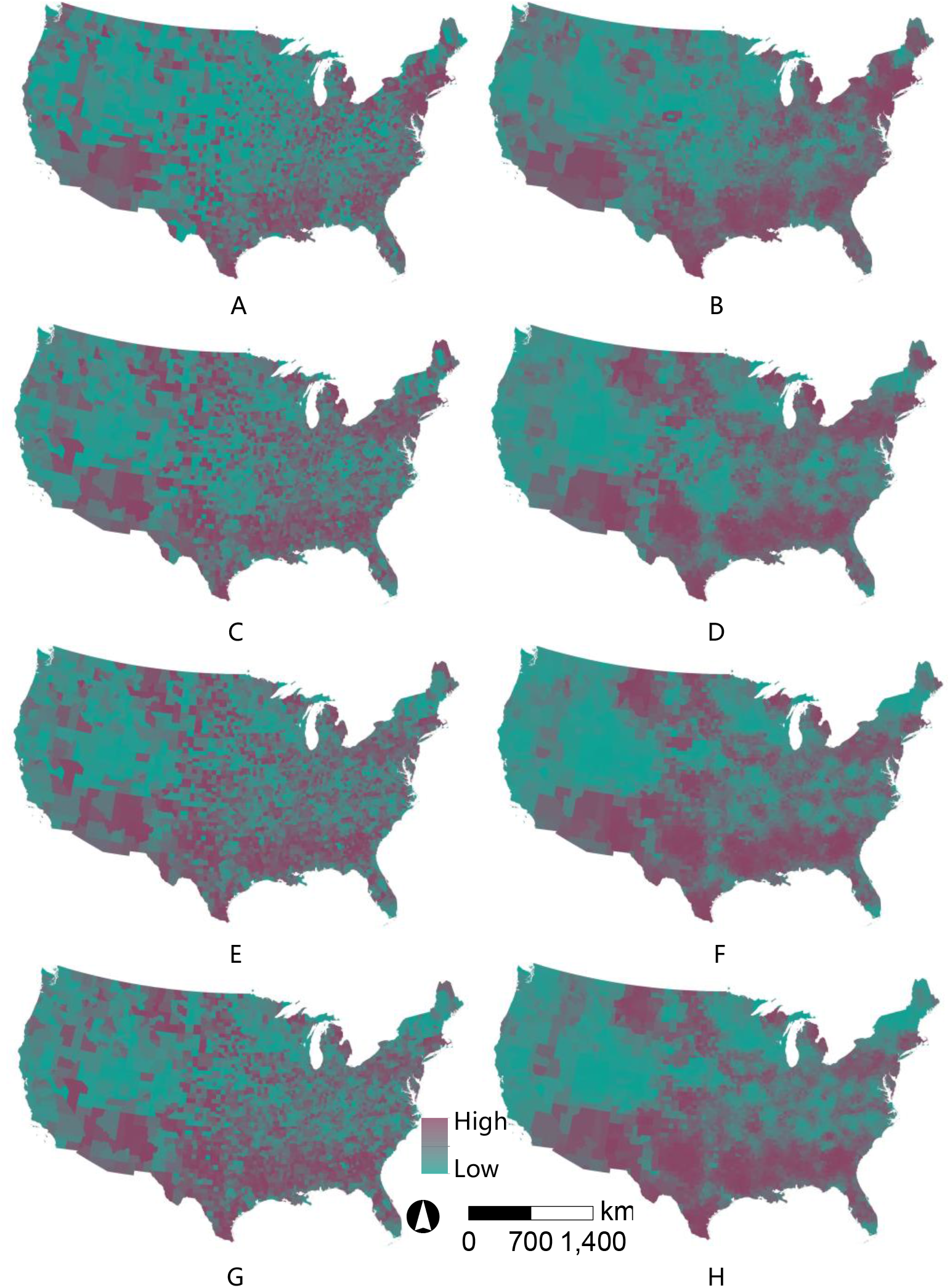
County-level COVID-19 fatality across the contiguous US Note: The figures on the left side from A to G show the distribution of fatality for period 1, period 2, period 3, and period 4, respectively. The corresponding right figures from B to H show the distribution of spatially lagged fatality for the corresponding time periods.

Figure 2 showed the geospatial distribution of Twitter-based place connectivity and concentrated disadvantage. It can be observed that high connected counties were like those major transportation nodes. The Northeast and Southwest were areas of higher place connectivity, which contained some notable metropolitan areas, such as San Francisco, Los Angeles, and New York City. The spatial distribution of place connectivity was similar in 2019 and 2018. The map of concentrated disadvantage showed that high disadvantaged counties were mainly located in the South, while coastal areas and some northern areas showed lower levels of disadvantage (Figure 2).

**Figure 2.**
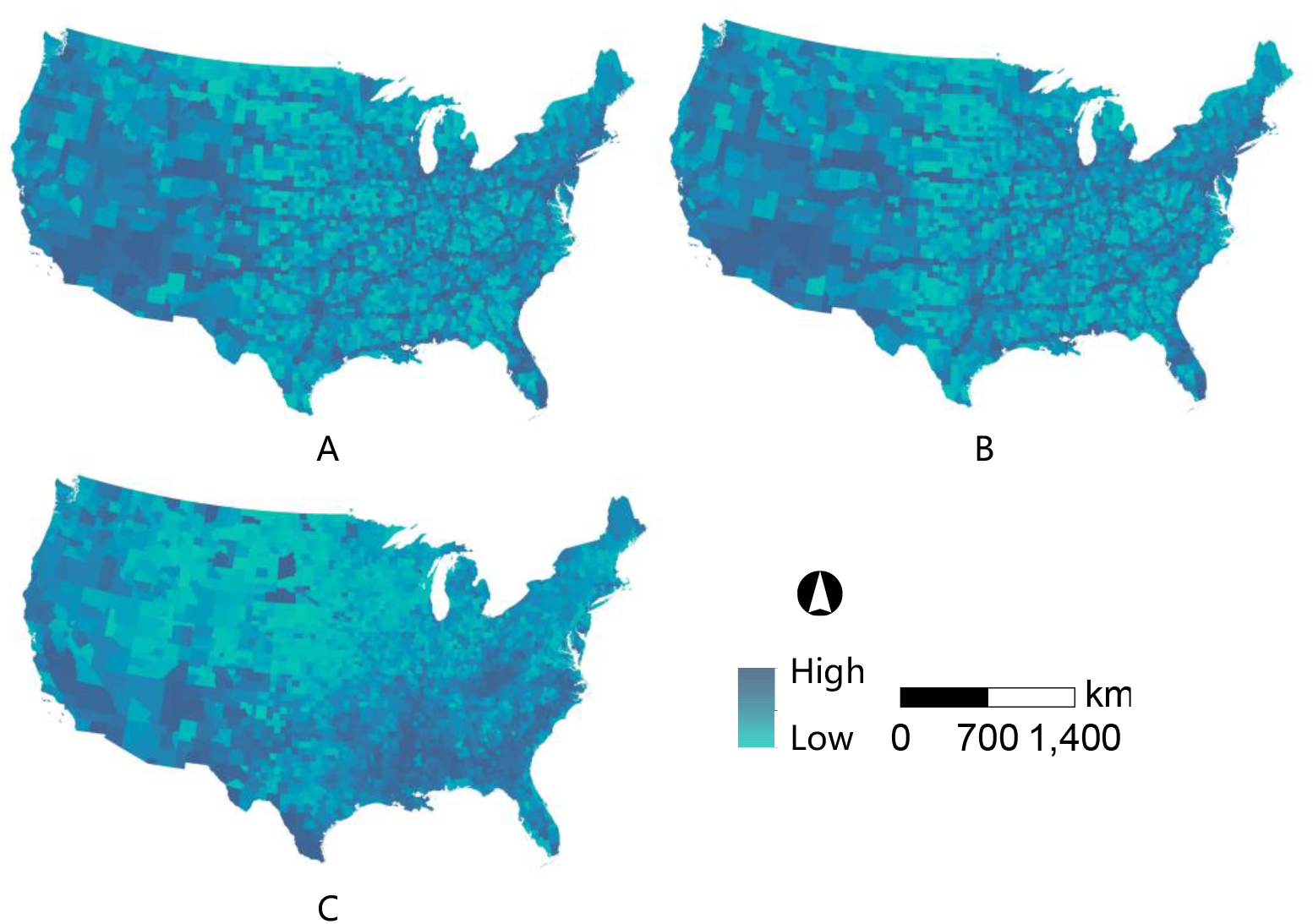
Twitter-based place connectivity across the contiguous US Note: A and B refer to place connectivity for 2019 and 2018, separately. C refers to the map of concentrated disadvantage.

### The results of mixed-effects negative binomial regression models

The results used the incidence rate ratio (IRR) to represent the association between the variables. An IRR greater than 1 represents a positive association between county-level factors and COVID-19 fatality. In contrast, an IRR less than 1 represents a negative association, and an IRR equal to 1 represents no positive or negative association. Model 1 in Tables 3 to 6 showed the results of mixed-effect negative binomial regression analysis up to the four-time points. In each period, counties with higher disadvantages had higher fatality than those with lower disadvantages (*IRR* > 1, *p* < 0.01). For example, in period 4, compared to counties with lower concentrated disadvantage, the fatality in counties with higher concentrated disadvantage was 1.157 times higher. (*IRR* = 1.157, 95% *CI*: 1.125-1.189, *p* < 0.01). It indicates hypothesis 1 is confirmed. The effects of place connectivity were significant in period 2, 3, & 4 (*IRR* <1, *p* < 0.01), but not in period 1(*IRR* < 1, *p* > 0.05). Specifically, counties with higher place connectivity had lower fatality than counties with lower place connectivity.

**Table 3.** Mixed-effects negative binomial regression models of county-level COVID-19 fatality (period 1)

| Factors | Model 1 | Model 2 |
| --- | --- | --- |
|  | IRR (95% *CI*) | IRR (95% *CI*) |
| Concentrated disadvantage | 1.185(1.119,1.255) ** | 1.188(1.121, 1.258) ** |
| Log (PC) | 0.93(0.812,1.065) | 0.908(0.791, 1.044) |
| Concentrated disadvantage * Log (PC) |  | 1.142(0.958, 1.361) |
| Log (Spatially lagged fatality) | 2.519(2.2,2.884) ** | 2.526(2.207, 2.892) ** |
| Log (population density) | 1.089(1.03,1.152) ** | 1.094(1.034, 1.157) ** |
| % of population aged 65 + | 1.039(1.033,1.046) ** | 1.039(1.033, 1.045) ** |
| % of no health insurance | 1(0.993,1.007) | 1.000(0.993, 1.007) |
| % of black or African Americans | 1.004(1.002,1.006) ** | 1.004(1.002, 1.006) ** |
| % of workers 16 years and over who commute by public transportation | 1.008(0.998,1.018) | 1.008(0.998, 1.018) |
| ICU beds per 100,000 population | 1(0.999,1.001) | 1(0.999, 1.001) |
| CBSA (Non-CBSA) |  |  |
| Micro | 0.963(0.9,1.03) | 0.961(0.898, 1.028) |
| Metro | 0.968(0.906,1.034) | 0.965(0.903, 1.031) |
| Region (Northeast) |  |  |
| Midwest | 0.616(0.511,0.742) ** | 0.614(0.510, 0.740) ** |
| South | 0.644(0.539,0.769) ** | 0.645(0.540, 0.770) ** |
| West | 0.62(0.507,0.758) ** | 0.619(0.507, 0.757) ** |

Notes: IRR: incidence rate ratio; PC: place connectivity; *CI*: Confidence interval; \*: $p < .05$ ; \*\*: $p < .001$ .

**Table 4.** Mixed-effects negative binomial regression models of county-level COVID-19 fatality (period 2)

| Factors | Model 1 | Model 2 |
| --- | --- | --- |
|  | IRR (95% *CI*) | IRR (95% *CI*) |
| Concentrated disadvantage | 1.156(1.117,1.197) ** | 1.166(1.126,1.206) ** |
| Log (PC) | 0.878(0.812,0.951) ** | 0.852(0.786,0.923) ** |
| Concentrated disadvantage * Log (PC) |  | 1.267(1.141,1.406) ** |
| Log (Spatially lagged fatality) | 2.799(2.467,3.176) ** | 2.795(2.464,3.169) ** |
| Log (population density) | 1.026(0.992,1.06) | 1.031(0.997,1.066) |
| % of population aged 65 + | 1.035(1.032,1.039) ** | 1.035(1.031,1.038) ** |
| % of no health insurance | 1.008(1.004,1.013) ** | 1.008(1.004,1.012) ** |
| % of black or African Americans | 1.002(1,1.003) ** | 1.002(1,1.003) * |
| % of workers 16 years and over who commute by public transportation | 1.005(0.999,1.011) | 1.005(0.999,1.011) |
| ICU beds per 100,000 population | 1(1,1.001) | 1(1,1.001) |
| CBSA (Non-CBSA) |  |  |
| Micro | 0.961(0.924,1) * | 0.958(0.921,0.996) * |
| Metro | 0.95(0.914,0.987) ** | 0.946(0.91,0.983) ** |
| Region (Northeast) |  |  |
| Midwest | 0.889(0.788,1.003) | 0.884(0.784,0.997) * |
| South | 0.822(0.731,0.923) ** | 0.824(0.734,0.925) ** |
| West | 0.792(0.696,0.901) ** | 0.79(0.694,0.899) ** |

Notes: IRR: incidence rate ratio; PC: place connectivity; *CI*: Confidence interval; \*: $p < .05$ ; \*\*: $p < .001$ .

**Table 5.** Mixed-effects negative binomial regression models of county-level COVID-19 fatality (period 3)

| Factors | Model 1 | Model 2 |
| --- | --- | --- |
|  | IRR (95% *CI*) | IRR (95% *CI*) |
| Concentrated disadvantage | 1.172(1.137,1.208) ** | 1.182(1.147,1.219) ** |
| Log (PC) | 0.885(0.826,0.948) ** | 0.854(0.796,0.916) ** |
| Concentrated disadvantage * Log (PC) |  | 1.303(1.189,1.428) ** |
| Log (Spatially lagged fatality) | 2.766(2.449,3.124) ** | 2.758(2.444,3.113) ** |
| Log (population density) | 1.016(0.987,1.046) | 1.022(0.992,1.052) |
| % of population aged 65 + | 1.035(1.032,1.039) ** | 1.034(1.031,1.038) ** |
| % of no health insurance | 1.008(1.004,1.012) ** | 1.008(1.004,1.011) ** |
| % of black or African Americans | 1.002(1.001,1.003) ** | 1.002(1.001,1.003) ** |
| % of workers 16 years and over who commute by public transportation | 1.004(0.999,1.009) | 1.004(0.999,1.009) |
| ICU beds per 100,000 population | 1(1,1.001) | 1(1,1.001) |
| CBSA (Non-CBSA) |  |  |
| Micro | 0.958(0.926,0.992) ** | 0.954(0.922,0.988) ** |
| Metro | 0.957(0.925,0.99) ** | 0.952(0.921,0.985) ** |
| Region (Northeast) |  |  |
| Midwest | 0.974(0.867,1.094) | 0.968(0.862,1.087) |
| South | 0.924(0.826,1.034) | 0.928(0.83,1.037) |
| West | 0.861(0.76,0.974) * | 0.859(0.759,0.971) * |

Notes: IRR: incidence rate ratio; PC: place connectivity; *CI*: Confidence interval; \*: $p < .05$ ; \*\*: $p < .001$ .

**Table 6.** Mixed-effects negative binomial regression models of county-level COVID-19 fatality (period 4)

| Factors | Model 1 | Model 2 |
| --- | --- | --- |
|  | IRR (95% *CI*) | IRR (95% *CI*) |
| Concentrated disadvantage | 1.157(1.125,1.189) ** | 1.167(1.135,1.199) ** |
| Log (PC) | 0.923(0.867,0.983) * | 0.888(0.834,0.946) ** |
| Concentrated disadvantage * Log (PC) |  | 1.321(1.216,1.436) ** |
| Log (Spatially lagged fatality) | 2.909(2.578,3.283) ** | 2.892(2.566,3.26) ** |
| Log (population density) | 1.009(0.983,1.036) | 1.015(0.989,1.042) |
| % of population aged 65 + | 1.033(1.03,1.036) ** | 1.032(1.029,1.035) ** |
| % of no health insurance | 1.008(1.004,1.011) ** | 1.008(1.004,1.011) ** |
| % of black or African Americans | 1.002(1.001,1.003) ** | 1.002(1.001,1.003) ** |
| % of workers 16 years and over who commute by public transportation | 1.003(0.999,1.008) | 1.003(0.999,1.008) |
| ICU beds per 100,000 population | 1(1,1.001) | 1(1,1.001) |
| CBSA (Non-CBSA) |  |  |
| Micro | 0.973(0.943,1.004) | 0.969(0.94,1) * |
| Metro | 0.968(0.939,0.998) * | 0.963(0.935,0.993) * |
| Region (Northeast) |  |  |
| Midwest | 0.978(0.876,1.092) | 0.973(0.872,1.086) |
| South | 0.951(0.856,1.058) | 0.955(0.859,1.062) |
| West | 0.901(0.802,1.012) | 0.899(0.801,1.009) |

Notes: IRR: incidence rate ratio; PC: place connectivity; *CI*: Confidence interval; \*: $p < .05$ ; \*\*: $p < .001$ .

Further, the results showed that the interaction between concentrated disadvantage and place connectivity was significant in three time periods (*IRR >* 1, *p* < 0.01), except for period 1(*IRR* > 1, *p* > 0.05), after the inclusion of an interaction term (Model 2, Tables 3 to 6). It supports hypothesis 2, implying that concentrated disadvantage is associated with a relative increase in the county-level COVID-19 fatality, especially for those counties with high place connectivity. Interestingly, the IRR of this interaction increased with the time periods, indicating an increasing robust interaction effect (Tables 3 to 6 and Figure 3), which confirms hypothesis 3. Figure 3 shows the graphical illustration of the interaction between concentrated disadvantage and place connectivity using results from Model 2, Tables 3 to 6. The relationship between concentrated disadvantage and fatality becomes stronger as the value of place connectivity increases.

**Figure 3.**
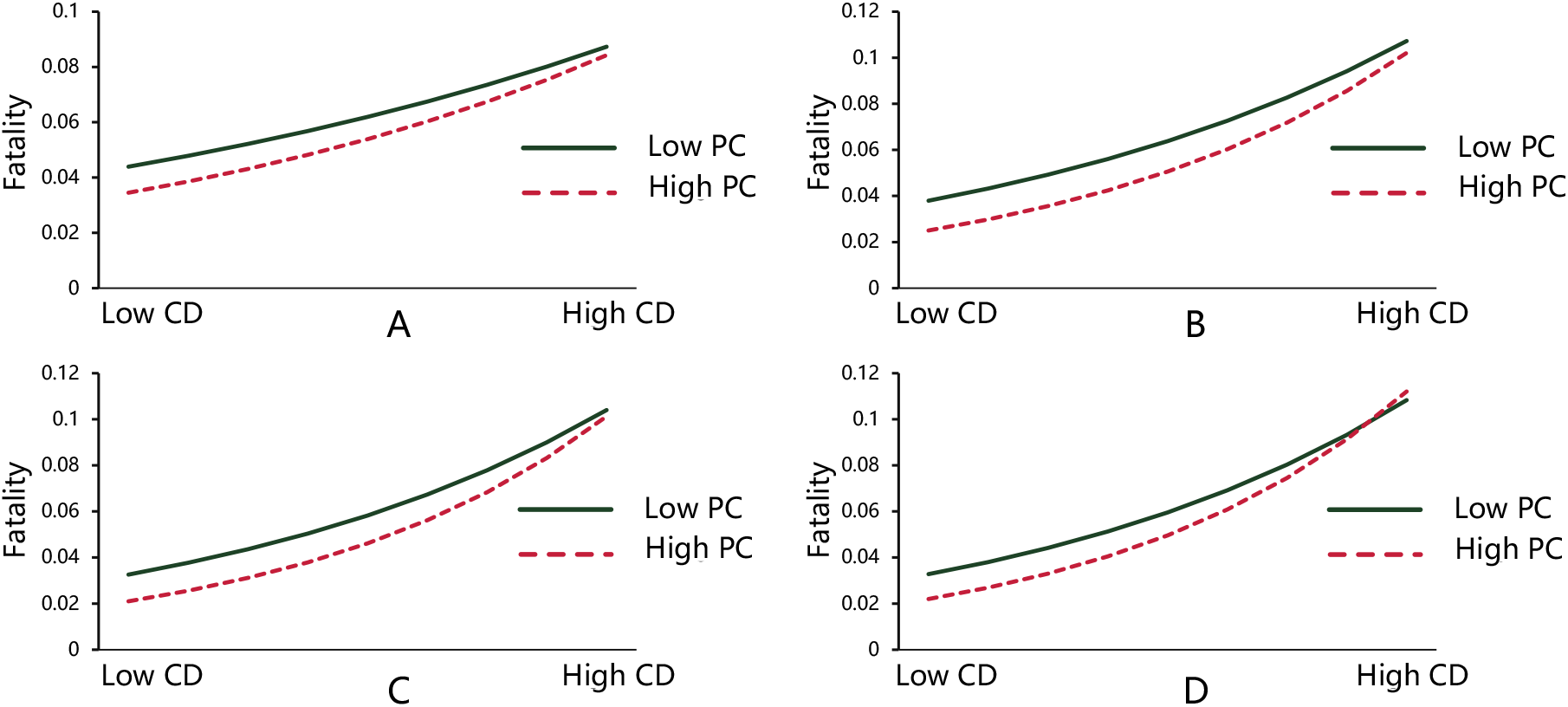
The impacts of concentrated disadvantage on COVID-19 fatality by different place connectivity Note: figures A to D refer to the models for time periods 1 to 4, respectively (the interaction for period 1 is not significant). CD refers to concentrated disadvantage, and PC refers to place connectivity.

Most control variables also showed consistent results across time (Model 1, Tables 3 to 6). Each 1 standard deviation increase in spatial lagged fatality rate was associated with a significant relative increase in the IRR of COVID-19 fatality (*IRR* > 1 and *p* < 0.01). A higher percentage of people aged 65 and older was associated with a higher COVID-19 fatality (*IRR* > 1 and *p* < 0.01). Similarly, a higher percentage of black or African Americans was associated with a higher COVID-19 fatality (*IRR* > 1 and *p* < 0.01). In addition, except for period 1, micro and metropolitan counties had lower COVID-19 fatality than rural counties (*IRR* < 1 and *p* < 0.01). The region factor was not consistently significant. Over time, the significant regional differences in COVID-19 fatality gradually disappeared. The remaining control variables did not show significant results.

We also ran the models using the 2018 place connectivity data, which showed consistent results to the model using 2019 place connectivity data (Tables 1 to 4 in Appendix).

## Discussion

Leveraging concentrated disadvantage and Twitter-based place connectivity, we examined the relationship between concentrated disadvantage and COVID-19 fatality in the US, and how this association is moderated by place connectivity. In addition to examining the harmful effect of concentrated disadvantage, this study partially explored the mechanism of this effect. The significant interaction between place connectivity and concentrated disadvantage suggests that socioeconomically disadvantaged groups in an area with high levels of place connectivity may be more likely to experience higher mobility, and thus face higher incidence and fatality risk. The results provide new insights into the association between concentrated disadvantage and COVID-19 fatality and may provide some guidance for future infectious disease control policies in socioeconomically disadvantaged areas.

We further found that the moderation effect of place connectivity increased over time, which may be related to increased mobility and the loosening of travel restrictions. At the early stages of the pandemic, COVID-19 fatality was more severe, travel restrictions were higher, and people were also in a precautionary awareness to reduce their outside activities, so the effect of concentrated disadvantage on fatality may be less influenced by place connectivity. In contrast, with widespread vaccination, people entered the New Normal, less restrictive in their travel, thus the moderation effect of place connectivity on the link between concentrated disadvantage and COVID-19 fatality became increasingly significant.

The significant association between place connectivity and decreased COVID-19 fatality rate is observed in time periods 2 to 4. We found a moderate correlation between population density, ICU percentage, and place connectivity. The high fatality was mostly found in districts with low population density due to poorer health care systems (Hamidi, Sabouri, & Ewing, 2020). Rural areas hold less access to health facilities, but urban areas, which are more likely to encounter large numbers of cases, instead have better health facility preparation and prevention to avoid more deaths (Ahmed et al., 2020). It implies that highly connected areas are generally areas with higher population density and urbanization, and may have better medical conditions and facilities, leading to a lower fatality. This finding is consistent with associations between urbanization (Ahmed et al., 2020; Iyanda, Boakye, & Oppong, 2020) and population density (Hamidi, Sabouri, & Ewing, 2020; Souris & Gonzalez, 2020), and lower COVID-19 fatality. However, the effect of place connectivity on COVID-19 was not significant in period 1, probably due to strict travel restrictions and low travel needs in the early stage, resulting in connectivity not working.

These findings have potential practical implications for the response to the COVID-19 pandemic. Socioeconomically disadvantaged areas with high place connectivity are likely to have higher population mobility. Reducing the degree of place connectivity, such as road, air transport, and other restrictions may reduce the population mobility of residents in disadvantaged areas, thus curbing infection and deaths among this population. However, these policies cannot eliminate people’s demands for daily commute (Sy et al., 2021). Policymakers need to balance the epidemic prevention and socioeconomic costs of travel restrictions. In future practice, we could pay extra attention to highly concentrated disadvantaged and highly connected areas and take additional measures for these areas to reduce disproportionate COVID-19 transmission and deaths, which could prevent the spread of the epidemic on a wider scale. Timely monitoring the epidemic of concentrated disadvantaged areas with high place connectivity and identifying potential epidemic hotpots and disadvantaged areas may assist evidence-based decision-making in resource allocation and tailored strategies to effectively respond to COVID-19 pandemic and other emerging infectious diseases in the future.

There are a few limitations to this study. First, place connectivity is measured from Twitter data, while is less used by some groups, such as the elderly and children. Also, data in some counties where Twitter is less used may be underrepresented. Second, utilizing county-level data to understand the effects of concentrated disadvantage on COVID-19 fatality may ignore the role of neighborhood-level factors. There may exist several neighborhoods with high socioeconomic status even in concentrated disadvantaged counties. Last, this study used spatial-scale variables rather than individual-level COVID-19 data, hence the results can only indicate the associations between geospatial environment and COVID-19 outcomes and cannot be interpreted as individual-level associations or causalities. Future multilevel analyses could be applied, including data on individual characteristics and neighborhood factors, which could yield more robust findings.

## Conclusion

Concentrated disadvantage contributes to the geospatial disparities in county-level COVID-19 fatality in the US: counties with higher levels of socioeconomic disadvantage reported higher levels of COVID-19 fatality. Place connectivity moderates the detrimental effects of concentrated disadvantage on fatality, and this moderation effect increases along with time periods. Our finding not only further explains the link between concentrated disadvantage and COVID-19 fatality, but also further highlights the role of place connectivity in combating COVID-19 and future infectious diseases. In response to COVID-19 and future infectious disease outbreaks, relevant policies may be specifically tailored for areas where socioeconomic disadvantage and high place connectivity coexist.

## Data Availability

All data produced are available online at

https://github.com/nytimes/covid-19-data

https://www.census.gov/programs-surveys/acs/data.html

## Appendix

**Table 1.**
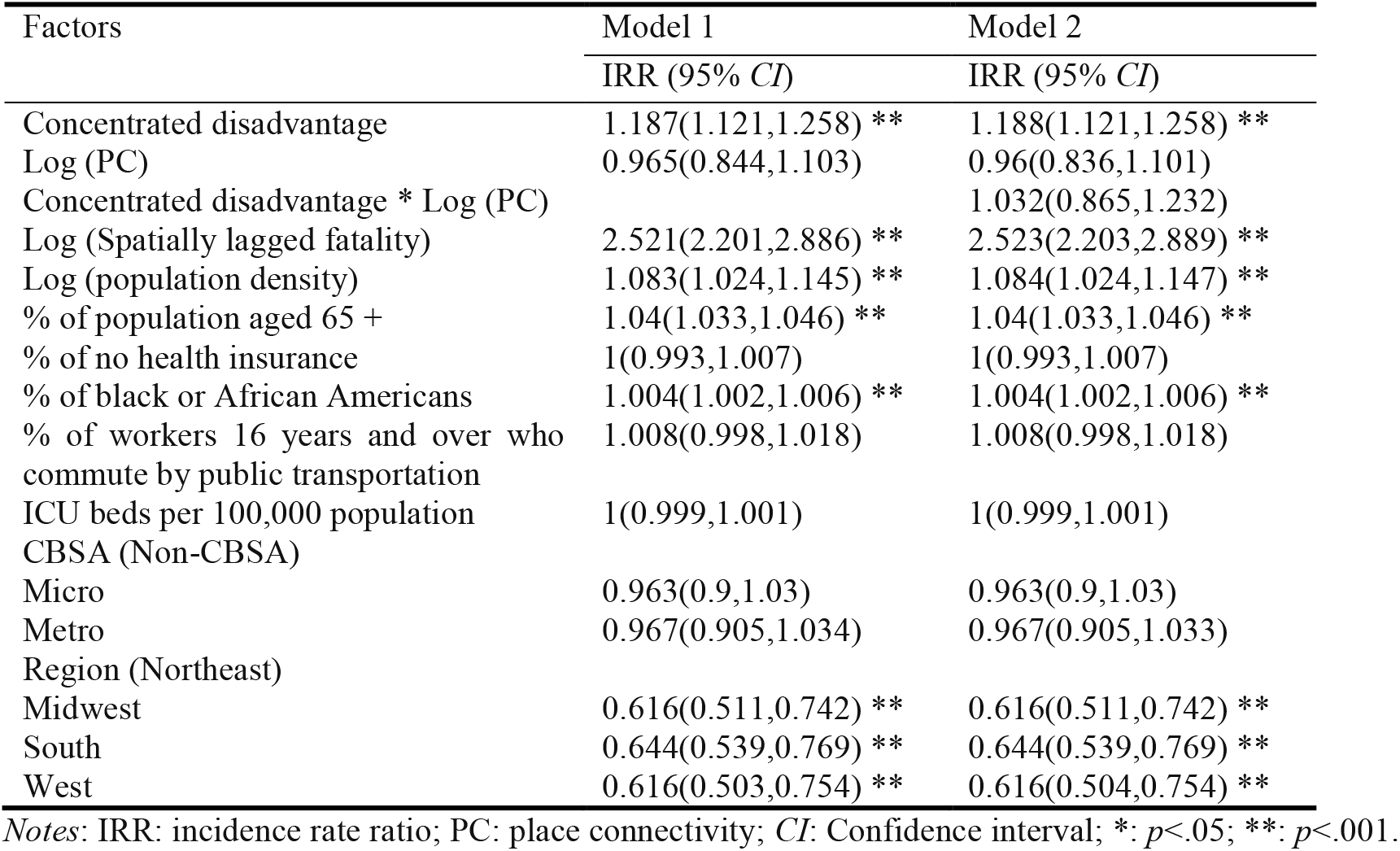
Mixed-effects negative binomial regression models of county-level COVID-19 fatality (period 1)

| Factors | Model 1 | Model 2 |
| --- | --- | --- |
|  | IRR (95% *CI*) | IRR (95% *CI*) |
| Concentrated disadvantage | 1.187(1.121,1.258) ** | 1.188(1.121,1.258) ** |
| Log (PC) | 0.965(0.844,1.103) | 0.96(0.836,1.101) |
| Concentrated disadvantage * Log (PC) |  | 1.032(0.865,1.232) |
| Log (Spatially lagged fatality) | 2.521(2.201,2.886) ** | 2.523(2.203,2.889) ** |
| Log (population density) | 1.083(1.024,1.145) ** | 1.084(1.024,1.147) ** |
| % of population aged 65 + | 1.04(1.033,1.046) ** | 1.04(1.033,1.046) ** |
| % of no health insurance | 1(0.993,1.007) | 1(0.993,1.007) |
| % of black or African Americans | 1.004(1.002,1.006) ** | 1.004(1.002,1.006) ** |
| % of workers 16 years and over who commute by public transportation | 1.008(0.998,1.018) | 1.008(0.998,1.018) |
| ICU beds per 100,000 population | 1(0.999,1.001) | 1(0.999,1.001) |
| CBSA (Non-CBSA) |  |  |
| Micro | 0.963(0.9,1.03) | 0.963(0.9,1.03) |
| Metro | 0.967(0.905,1.034) | 0.967(0.905,1.033) |
| Region (Northeast) |  |  |
| Midwest | 0.616(0.511,0.742) ** | 0.616(0.511,0.742) ** |
| South | 0.644(0.539,0.769) ** | 0.644(0.539,0.769) ** |
| West | 0.616(0.503,0.754) ** | 0.616(0.504,0.754) ** |

Notes: IRR: incidence rate ratio; PC: place connectivity; *CI*: Confidence interval; \*: $p < .05$ ; \*\*: $p < .001$ .

**Table 2.**
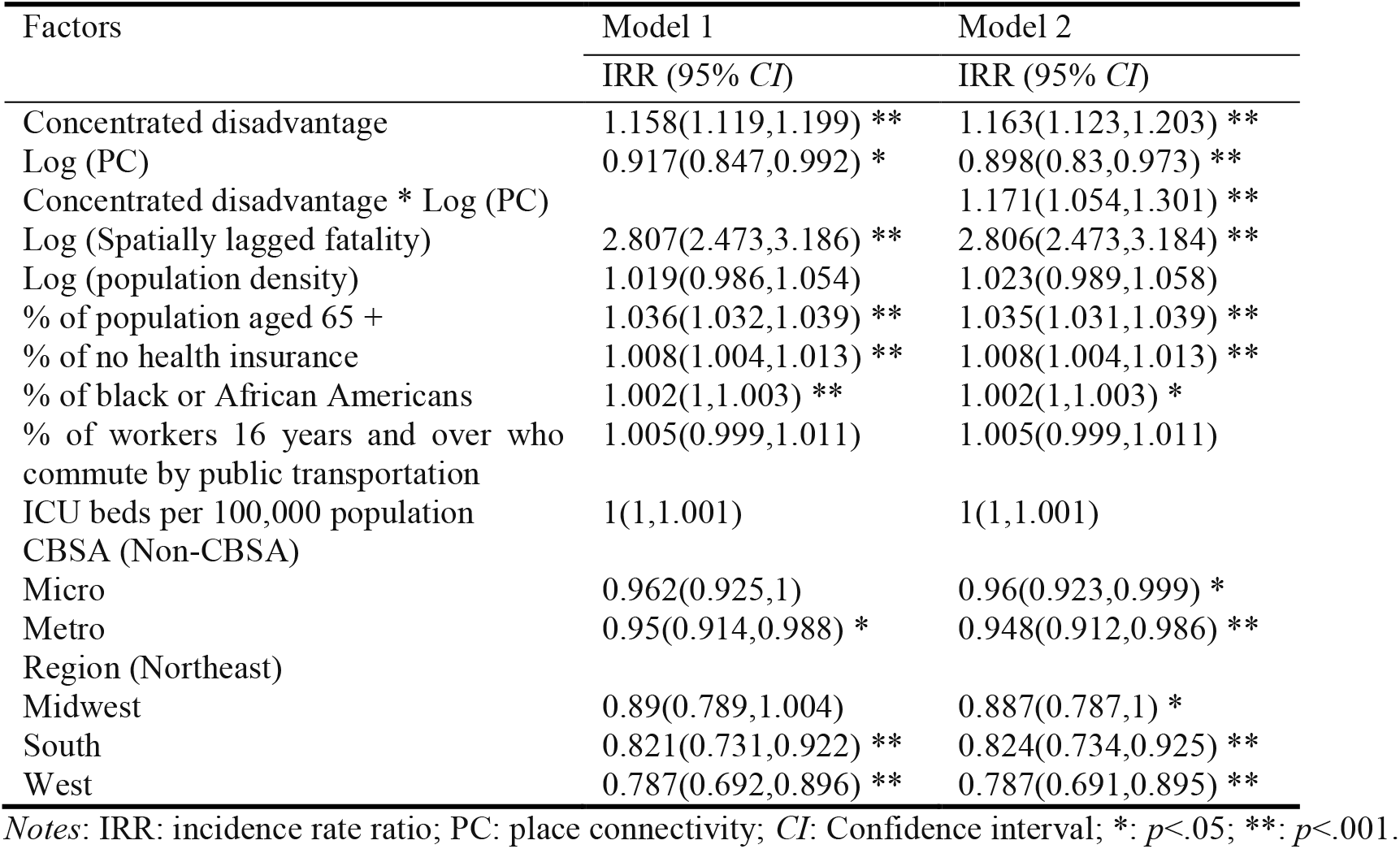
Mixed-effects negative binomial regression models of county-level COVID-19 fatality (period 2)

**Table 3.**
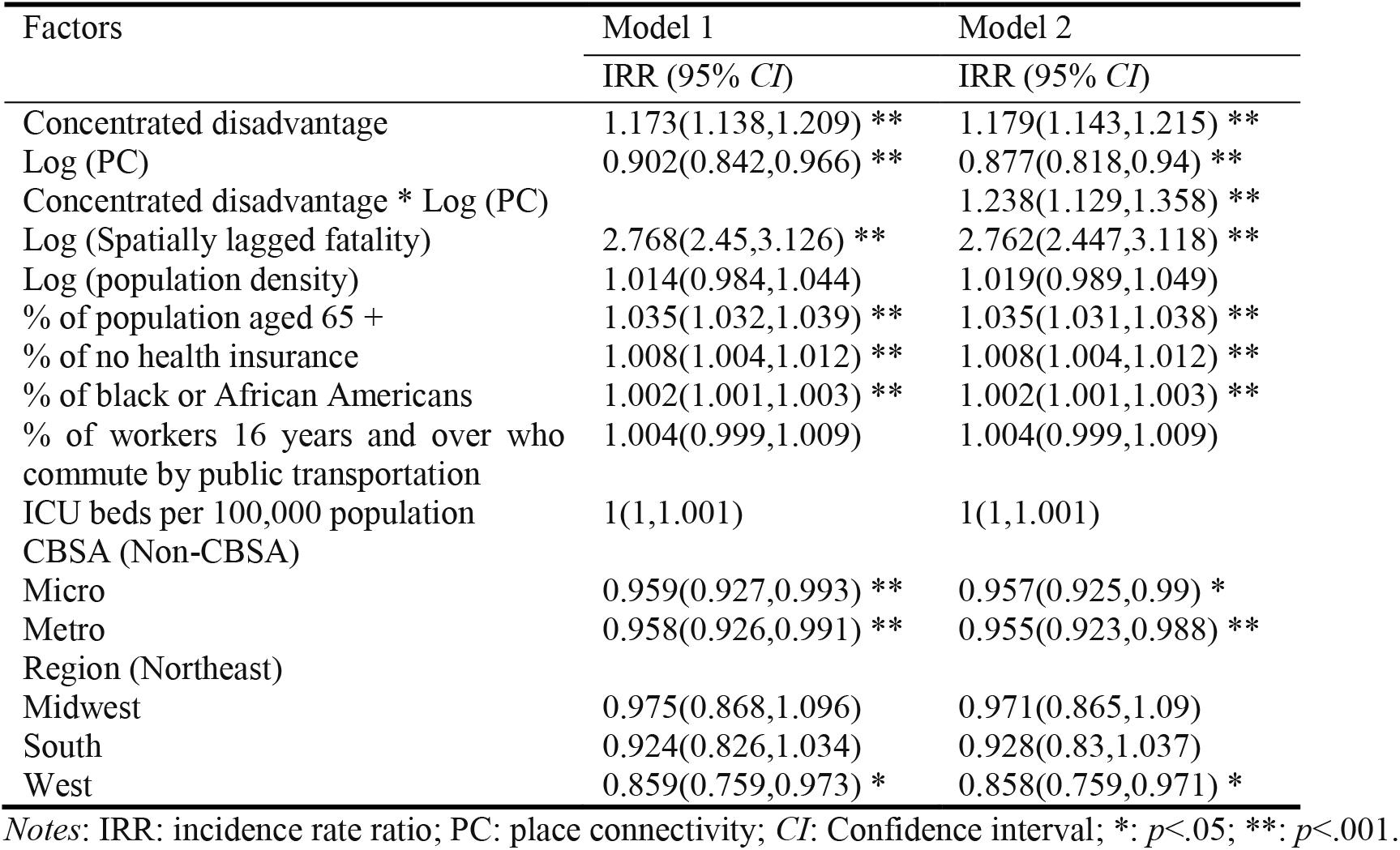
Mixed-effects negative binomial regression models of county-level COVID-19 fatality (period 3)

**Table 4.** Mixed-effects negative binomial regression models of county-level COVID-19 fatality (period 4)

| Factors | Model 1 | Model 2 |
| --- | --- | --- |
|  | IRR (95% *CI*) | IRR (95% *CI*) |
| Concentrated disadvantage | 1.158(1.126,1.19) * | 1.164(1.132,1.196) ** |
| Log (PC) | 0.942(0.885,1.002) | 0.913(0.858,0.972) ** |
| Concentrated disadvantage * Log (PC) |  | 1.248(1.148,1.357) ** |
| Log (Spatially lagged fatality) | 2.908(2.577,3.282) ** | 2.889(2.562,3.258) ** |
| Log (population density) | 1.006(0.98,1.033) | 1.011(0.985,1.039) |
| % of population aged 65 + | 1.033(1.03,1.036) ** | 1.032(1.029,1.035) ** |
| % of no health insurance | 1.008(1.005,1.012) ** | 1.008(1.004,1.011) ** |
| % of black or African Americans | 1.002(1.001,1.003) ** | 1.002(1.001,1.003) ** |
| % of workers 16 years and over who commute by public transportation | 1.003(0.999,1.008) | 1.003(0.998,1.008) |
| ICU beds per 100,000 population | 1(1,1.001) | 1(1,1.001) |
| CBSA (Non-CBSA) |  |  |
| Micro | 0.974(0.944,1.004) | 0.971(0.942,1.002) |
| Metro | 0.969(0.94,0.999) * | 0.966(0.937,0.995) * |
| Region (Northeast) |  |  |
| Midwest | 0.979(0.877,1.093) | 0.975(0.873,1.088) |
| South | 0.951(0.855,1.057) | 0.956(0.86,1.062) |
| West | 0.899(0.8,1.009) | 0.898(0.8,1.008) |

Notes: IRR: incidence rate ratio; PC: place connectivity; *CI*: Confidence interval; \*: $p < .05$ ; \*\*: $p < .001$ .

## References

Adie, Y., Kats, D. J., Tlimat, A., Perzynski, A., Dalton, J., Gunzler, D., & Tarabichi, Y. (2020). Neighborhood disadvantage and lung cancer incidence in ever-smokers at a safety net health-care system: a retrospective study. Chest, 157(4), 1021–1029.

Ahmed, R., Williamson, M., Hamid, M. A., & Ashraf, N. (2020, September). United States county-level COVID-19 death rates and case fatality rates vary by region and urban status. In Healthcare (Vol. 8, No. 3, p. 330). Multidisciplinary Digital Publishing Institute.

Barber, L. E., Zirpoli, G. R., Cozier, Y. C., Rosenberg, L., Petrick, J. L., Bertrand, K. A., & Palmer, J. R. (2021). Neighborhood disadvantage and individual-level life stressors in relation to breast cancer incidence in US Black women. Breast Cancer Research, 23(1), 1–12.

Benitez, J., Courtemanche, C., & Yelowitz, A. (2020). Racial and ethnic disparities in COVID-19: evidence from six large cities. Journal of Economics, Race, and Policy, 3(4), 243–261.

Boateng, Godfred O., Laura M. Phipps, Laura E. Smith, and Frederick A. Armah. “Household energy insecurity and COVID-19 have independent and synergistic health effects on vulnerable populations.” Frontiers in Public Health 8 (2021): 1068.

Bonaccorsi, G., Pierri, F., Cinelli, M., Flori, A., Galeazzi, A., Porcelli, F., … & Pammolli, F. (2020). Economic and social consequences of human mobility restrictions under COVID-19. Proceedings of the National Academy of Sciences, 117(27), 15530–15535.

Correa-Agudelo, E., Mersha, T. B., Branscum, A. J., MacKinnon, N. J., & Cuadros, D. F. (2021). Identification of vulnerable populations and areas at higher risk of Covid-19-related mortality during the early stage of the epidemic in the United States. International journal of environmental research and public health, 18(8), 4021.

Cuadros, D. F., Xiao, Y., Mukandavire, Z., Correa-Agudelo, E., Hernández, A., Kim, H., & MacKinnon, N. J. (2020). Spatiotemporal transmission dynamics of the COVID-19 pandemic and its impact on critical healthcare capacity. Health & place, 64, 102404.

DeGuzman, P. B., Cohn, W. F., Camacho, F., Edwards, B. L., Sturz, V. N., & Schroen, A. T. (2017). Impact of urban neighborhood disadvantage on late stage breast cancer diagnosis in Virginia. Journal of Urban Health, 94(2), 199–210.

Duque, R. B. (2021). Black health matters too… especially in the era of Covid-19: how poverty and race converge to reduce access to quality housing, safe neighborhoods, and health and wellness services and increase the risk of co-morbidities associated with global pandemics. Journal of Racial and Ethnic Health Disparities, 8(4), 1012–1025.

Durfey, S. N., Kind, A. J., Buckingham, W. R., DuGoff, E. H., & Trivedi, A. N. (2019). Neighborhood disadvantage and chronic disease management. Health services research, 54, 206–216.

Fountoulakis, K. N., Fountoulakis, N. K., Koupidis, S. A., & Prezerakos, P. E. (2020). Factors determining different death rates because of the COVID-19 outbreak among countries. Journal of Public Health, 42(4), 681–687.

Ghilarducci, T., & Farmand, A. (2020). Older workers on the COVID-19-frontlines without paid sick leave. Journal of Aging & Social Policy, 32(i4-5), 471–476.

Gupta, S., Kumar Patel, K., Sivaraman, S., & Mangal, A. (2020). Global epidemiology of first 90 days into COVID-19 pandemic: disease incidence, prevalence, case fatality rate and their association with population density, urbanisation and elderly population. Journal of Health Management, 22(2), 117–128.

Hamidi, S., Sabouri, S., & Ewing, R. (2020). Does density aggravate the COVID-19 pandemic? Early findings and lessons for planners. Journal of the American Planning Association, 86(4), 495–509.

Holmes, L., Enwere, M., Williams, J., Ogundele, B., Chavan, P., Piccoli, T., … & Dabney, K. W. (2020). Black–White risk differentials in COVID-19 (SARS-COV2) transmission, mortality and case fatality in the United States: translational epidemiologic perspective and challenges. International journal of environmental research and public health, 17(12), 4322.

Huyser, K. R., Yang, T. C., & Horse, A. J. Y. (2021). Indigenous Peoples, concentrated disadvantage, and income inequality in New Mexico: a ZIP code-level investigation of spatially varying associations between socioeconomic disadvantages and confirmed COVID-19 cases. J Epidemiol Community Health, 75(11), 1044–1049.

Iyanda, A. E., Boakye, K. A., Lu, Y., & Oppong, J. R. (2022). Racial/ethnic heterogeneity and rural-urban disparity of COVID-19 case fatality ratio in the USA: a negative binomial and GIS-based analysis. Journal of Racial and Ethnic Health Disparities, 9(2), 708–721.

Janke, Alexander T., Hao Mei, Craig Rothenberg, Robert D. Becher, Zhenqiu Lin, and Arjun K. Venkatesh. “Analysis of hospital resource availability and COVID-19 mortality across the United States.” Journal of hospital medicine 16, no. 4 (2021): 211–214.

Jr, C. A. (2021). Neighborhood disadvantage measures and COVID-19 cases in Boston, 2020. Public Health Reports, 136(3), 368–374.

Khanijahani, A., & Tomassoni, L. (2022). Socioeconomic and racial segregation and COVID-19: concentrated disadvantage and black concentration in association with COVID-19 deaths in the USA. Journal of racial and ethnic health disparities, 9(1), 367–375.

Khazanchi, R., Beiter, E. R., Gondi, S., Beckman, A. L., Bilinski, A., & Ganguli, I. (2020). County-level association of social vulnerability with COVID-19 cases and deaths in the USA. Journal of general internal medicine, 35(9), 2784–2787.

Kim, J. (2010). Neighborhood disadvantage and mental health: The role of neighborhood disorder and social relationships. Social science research, 39(2), 260–271.

Kirby, J. B., & Kaneda, T. (2005). Neighborhood socioeconomic disadvantage and access to health care. Journal of health and social behavior, 46(1), 15–31.

Kranjac, A. W., & Kranjac, D. (2021). County-level factors that influenced the trajectory of COVID-19 incidence in the New York City area. Health security, 19(S1), S–27.

Lee, M. R., Maume, M. O., & Ousey, G. C. (2003). Social isolation and lethal violence across the metro/nonmetro divide: The effects of socioeconomic disadvantage and poverty concentration on homicide. Rural sociology, 68(1), 107–131.

Lee, W., Kim, H., Choi, H. M., Heo, S., Fong, K. C., Yang, J., … & Bell, M. L. (2021). Urban environments and COVID-19 in three Eastern states of the United States. Science of The Total Environment, 779, 146334.

Levy, B. L., Vachuska, K., Subramanian, S. V., & Sampson, R. J. (2022). Neighborhood socioeconomic inequality based on everyday mobility predicts COVID-19 infection in San Francisco, Seattle, and Wisconsin. Science advances, 8(7), eabl3825.

Li, Z., Huang, X., Ye, X., Jiang, Y., Martin, Y., Ning, H., … & Li, X. (2021). Measuring global multi-scale place connectivity using geotagged social media data. Scientific reports, 11(1), 1–19.

Moosa, I. A., & Khatatbeh, I. N. (2021). Robust and fragile determinants of the infection and case fatality rates of Covid-19: international cross-sectional evidence. Applied Economics, 53(11), 1225–1234.

Peters, D. H., Garg, A., Bloom, G., Walker, D. G., Brieger, W. R., & Hafizur Rahman, M. (2008). Poverty and access to health care in developing countries. Annals of the new York Academy of Sciences, 1136(1), 161–171.

Pierce, J. B., Harrington, K., McCabe, M. E., Petito, L. C., Kershaw, K. N., Pool, L. R., … & Khan, S. S. (2021). Racial/ethnic minority and neighborhood disadvantage leads to disproportionate mortality burden and years of potential life lost due to COVID-19 in Chicago, Illinois. Health & place, 68, 102540.

Ross, C. E., & Mirowsky, J. (2001). Neighborhood disadvantage, disorder, and health. Journal of health and social behavior, 258–276.

Sampson, R. J., Raudenbush, S. W., & Earls, F. (1997). Neighborhoods and violent crime: A multilevel study of collective efficacy. science, 277(5328), 918–924.

Sampson, R. J., Sharkey, P., & Raudenbush, S. W. (2008). Durable effects of concentrated disadvantage on verbal ability among African-American children. Proceedings of the National Academy of Sciences, 105(3), 845–852.

Samuels-Kalow, M. E., Dorner, S., Cash, R. E., Dutta, S., White, B., Ciccolo, G. E., … & Camargo Jr, C. A. (2021). Neighborhood disadvantage measures and COVID-19 cases in Boston, 2020. Public Health Reports, 136(3), 368–374.

Sen-Crowe, B., Lin, I. C., Alfaro, R., McKenney, M., & Elkbuli, A. (2021). COVID-19 fatalities by zip codes and socioeconomic indicators across various US regions. Annals of Medicine and Surgery, 67, 102471.

Sen-Crowe, B., Sutherland, M., McKenney, M., & Elkbuli, A. (2021). A closer look into global hospital beds capacity and resource shortages during the COVID-19 pandemic. journal of Surgical Research, 260, 56–63.

Souris, M., & Gonzalez, J. P. (2020). COVID-19: Spatial analysis of hospital case-fatality rate in France. PLoS One, 15(12), e0243606.

Sun, X., Wandelt, S., & Zhang, A. (2021). On the degree of synchronization between air transport connectivity and COVID-19 cases at worldwide level. Transport Policy, 105, 115–123.

Sy, K. T. L., Martinez, M. E., Rader, B., & White, L. F. (2021). Socioeconomic disparities in subway use and COVID-19 outcomes in New York City. American Journal of Epidemiology, 190(7), 1234–1242.

Tokey, A. I. (2021). Spatial association of mobility and COVID-19 infection rate in the USA: A county-level study using mobile phone location data. Journal of Transport & Health, 22, 101135.

Truong, N., & Asare, A. O. (2021). Assessing the effect of socio-economic features of low-income communities and COVID-19 related cases: An empirical study of New York City. Global Public Health, 16(1), 1–16.

Wali, B., & Frank, L. D. (2021). Neighborhood-level COVID-19 hospitalizations and mortality relationships with built environment, active and sedentary travel. Health & Place, 71, 102659.

Wang, F., & Arnold, M. T. (2008). Localized income inequality, concentrated disadvantage and homicide. Applied Geography, 28(4), 259–270.

Yellow Horse, A. J., Yang, T. C., & Huyser, K. R. (2022). Structural inequalities established the architecture for COVID-19 pandemic among native Americans in Arizona: a geographically weighted regression perspective. Journal of Racial and Ethnic Health Disparities, 9(1), 165–175.

Zeng, C., Zhang, J., Li, Z., Sun, X., Yang, X., Olatosi, B., … & Li, X. (2021, October). Population mobility and health disparities in COVID-19 outbreaks in deep south: A county-level longitudinal analysis. In APHA 2021 Annual Meeting and Expo. APHA.

Zhang, Y., Zhang, A., & Wang, J. (2020). Exploring the roles of high-speed train, air and coach services in the spread of COVID-19 in China. Transport Policy, 94, 34–42.

Zhu, P., & Guo, Y. (2021). The role of high-speed rail and air travel in the spread of COVID-19 in China. Travel medicine and infectious disease, 42, 102097.

